# Implementing Primary Care Mediated Population Genetic Screening within an Integrated Health System

**DOI:** 10.1101/2020.07.16.20140228

**Authors:** Sean P David, Henry M Dunnenberger, Raabiah Ali, Adam Matsil, Amy A Lemke, Lavisha Singh, Anjali Zimmer, Peter J Hulick

**Affiliations:** NorthShore University HealthSystem, Evanston, IL, USA; Pritzker School of Medicine, The University of Chicago, Chicago, IL, USA; Color Genomics, Inc., Burlingame, CA, USA

## Abstract

**Introduction:** Genetic screenings can have a large impact on enabling personalized preventative care. However, this can be limited by the primary use of medical history-based screenings in determining care. The purpose of this study was to understand the impact of DNA10K, a population-based genetic screening program mediated by primary care physicians (PCPs) within an integrated health system to emphasize its contribution to preventative healthcare.

**Methods:** Construction of the patient experience as part of DNA10K shaped the context for PCP engagement within the program. A cross-sectional analysis of patient consents, orders, tests, and results of nearly 10,000 patients within the primary care specialties of family medicine, internal medicine or obstetrics/gynecology between April 1, 2019 and January 22, 2020 was conducted.

**Results:** Across all specialties, a median number of 7.5 cancer and cardiovascular disease variants per PCP was found. The average age of the study population was 49.6 years. Over 8% of these patients had at least one actionable genetic risk variant and almost 2% of patients had at least one CDC Tier 1 variant. The median number of patients per PCP with either hereditary breast and ovarian cancer, Lynch Syndrome, or Familial Hypercholesterolemia was 1 (Interquartile Range 0-2).

**Discussion:** The analysis of test results and the engagement of an integrated healthcare system in the implementation of a genetic screening program suggests that it can have a large impact on population health outcomes and minimal referral burden to PCPs if identified risks can lead to preventative care.

## Introduction

Genetic testing for pathogenic variants that confer susceptibility for cancer and heart disease – the two most common causes of death in the United States – has potential to guide individualized disease prevention, treatment, and risk identification. Primary care physicians (PCPs) play a pivotal role in preventive services, such as applying family histories in identifying high genetic risk for disease. However, medical history-based screening can under-detect pathogenic cancer syndromes such as Hereditary Breast and Ovarian Cancer (HBOC), Lynch Syndrome, and Familial Hypercholesterolemia (FH).^1-3^ The purpose of this study was to characterize the impact of PCP engagement in a population genetic screening program (DNA10K) by analyzing patient participation and test results.

## Methods

The DNA10K program was carried out within an integrated healthcare system to provide genomics-guided care to more than 10,000 patients by their PCPs (N=116) in family medicine (FM), internal medicine (IM), and obstetrics/gynecology (OBGYN) across 14 clinical practice locations in northern Cook and Lake Counties, Illinois. The context of the DNA10K implementation is described in Figure 1 [insert Figure 1].

**Figure 1.**
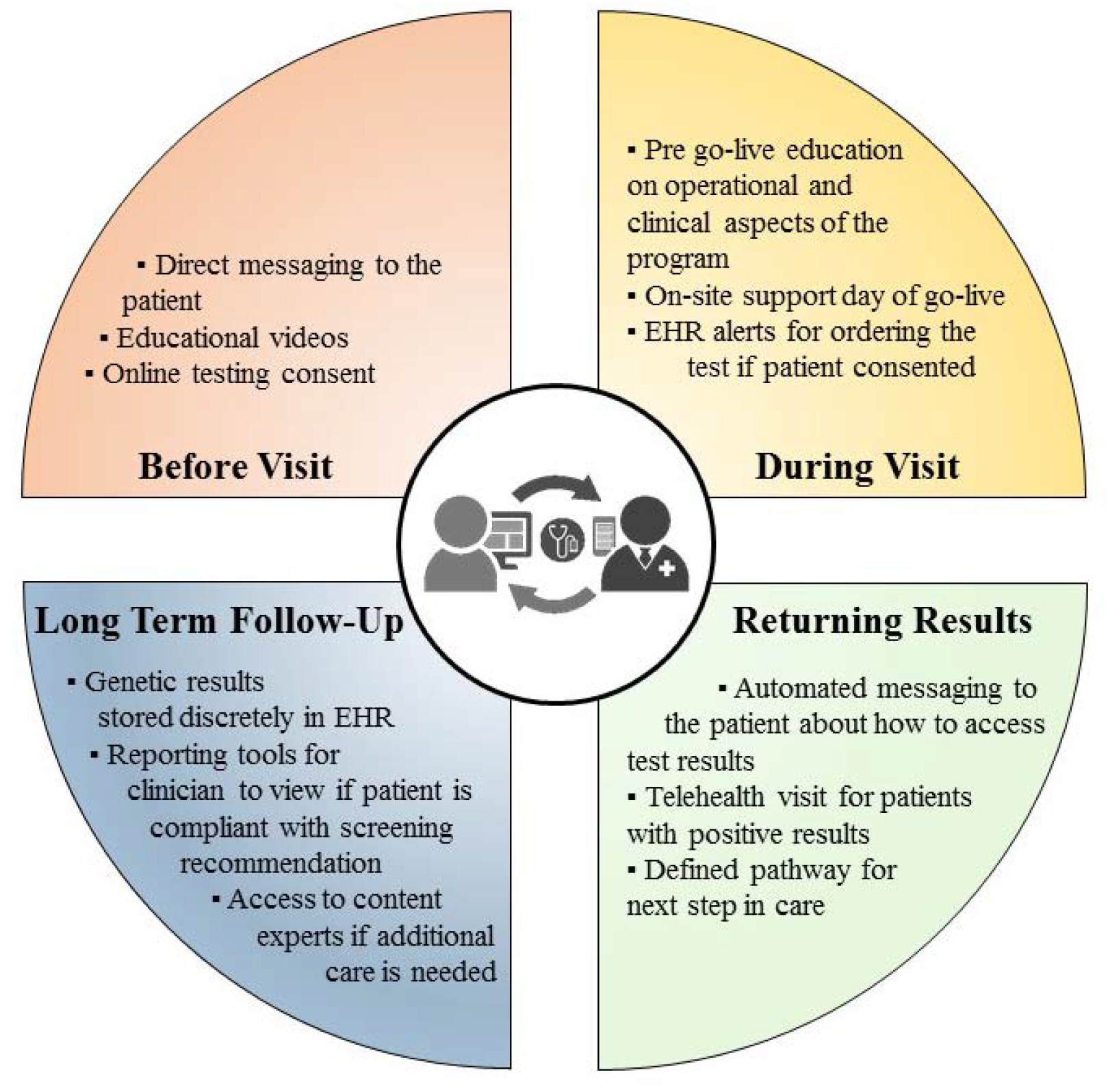
Context for Supporting PCP Engagement in Population Genetic Screening. The NorthShore Center for Personalized Medicine DNA10K program consisted of multiple components – (a) patient educational video and consent via the NorthShore Connect (NSC) patient portal, (b) PCP electronic clinical decision support and electronic medical record (EMR) order, (c) access to a NorthShore lab for phlebotomy, (d) Color Genomics, Inc. genetic testing using a 74 gene next-generation sequencing panel, (e) discrete reporting of results available in the EHR, (f) coordination of results with PCP health maintenance visits, (g) automated patient messaging about next steps, (h) access to genetic counseling, (i) return of results on the patient portal, and (j) clinical follow-up with PCP and/or specialists. This study was approved by the NorthShore Institutional Review Board as exempt quality improvement evaluation.

A cross-sectional analysis of consents, orders, tests, results, and actionable genetic variants of patients who were genotyped between April 1, 2019 and January 22, 2020 was conducted. Medians and interquartile ranges of genetic test result frequencies per PCP are reported at the overall and specialty levels for cancer and cardiovascular pathogenic variants, Tier 1 pathogenic variants^1^, non-Tier 1 pathogenic variants, and the percentage of actionable pharmacogenomic variants.^4^ Group differences for demographics and test results were examined using Kruskal Wallis, ANOVA with post-hoc Tukey HSD, and Wilcoxon Rank Sum tests in Statistical Analysis System 9.4 (p<0.05 considered significant).

## Results

Overall, 49,413 patients were contacted. Of those who consented (N=14,063), 77.7% (N=10,933) had an order placed by a PCP and 89.6% (N=9,797) of these patients completed testing. Mean age of the study population was 49.6 years. An additional 442 patients completed testing ordered by medical geneticists and other specialists (DNA10K total N=10,239). Table 1 presents stratified results by primary care specialty for DNA10K patient participation. A total of 813 (8.3%) patients had at least one actionable genetic risk variant (not including pharmacogenomics), and 182 (1.9%) patients had at least one CDC Tier 1 variant. HBOC variants were present in 116 patients (1.2%), Lynch Syndrome in 38 (0.39%) patients, and FH in 29 (0.3%) patients. More than 99% of patients (N=9,599/9,607) had at least one actionable pharmacogenomic variant. The median number of tested patients per PCP ranged from 62.5 (IQR 33-120) for FM, 66 (IQR 22.5-138) for IM, and 75 (IQR 40-118.2) for OBGYN. The median numbers of patients per PCP with at least one CDC Tier 1 variant or at least one actionable cancer or cardiovascular disease variant is 1 (IQR 0-2) and 5 (IQR 1-10), respectively. No statistically significant differences were observed per specialty PCP for test results analyzed in this analysis. [insert Table 1].

**Table 1.** DNA10K Primary Care Physician Mediated Genetic Screening Results.

| Variable | Overall<br>No./Total (%) or<br>Mean ± SD or<br>Median (IQR) | FM<br>No./Total (%)<br>or Median<br>(IQR) | IM<br>No./Total (%)<br>or Median<br>(IQR) | OBGYN<br>No./Total (%)<br>or Median<br>(IQR) | p-value<sup>a</sup> |
| --- | --- | --- | --- | --- | --- |
| **Demographics** |  |  |  |  |  |
| PCPs | 116 | 28 | 72 | 16 | ... |
| Patients Contacted | 49413 | 10640 | 32220 | 6553 | ... |
| Tests Consented | 14063/49419<br>(28.5%) | 3143/10640<br>(29.5%) | 8956/32220<br>(27.8%) | 1964/6553<br>(30.0%) | 0.13 |
| Orders Placed | 10933/14063<br>(77.7%) | 2416/3143<br>(76.9%) | 6920/8956<br>(77.3%) | 1597/1964<br>(81.3%) | 0.60 |
| Test Completed | 9797/10933<br>(89.6%) | 2190/2416<br>(90.6%) | 6255/6920<br>(90.4%) | 1352/1597<br>(84.7%) | 0.24 |
| **Patient Demographics** |  |  |  |  |  |
| Age | 49.6 ± 6.4 | 46.5 ± 4.9 | 52.3 ± 4.9 | 40.5 ± 3.3 | **<0.0001**<sup>b,c,d,e</sup> |
| Female | 6464/9797<br>(66.0%) | 1270/2190<br>(58.0%) | 3844/6255<br>(61.5%) | 1350/1352<br>(99.9%) | 0.80<sup>f</sup> |
| African American | 272/9797 (2.8%) | 100/2190 (4.6%) | 108/6255 (1.7%) | 64/1352 (4.7%) | **0.02**<sup>c</sup> |
| American<br>Indian/Alaskan Native | 22/9797 (0.22%) | 6/2190 (0.27%) | 14/6255 (0.22%) | 2/1352 (0.15%) | 0.86 |
| Asian | 679/9797 (6.9%) | 234/2190<br>(10.7%) | 368/6255 (5.8%) | 77/1352 (5.7%) | **0.04**<sup>d</sup> |
| Caucasian | 6528/9797<br>(66.6%) | 1229/2190<br>(56.1%) | 4424/6255<br>(70.7%) | 875/1352<br>(64.7%) | **0.04**<sup>c</sup> |
| Hispanic/Latino | 461/9797 (4.7%) | 145/2190 (6.6%) | 218/6255 (3.5%) | 98/1352 (7.2%) | **0.01**<sup>c,e</sup> |
| Pacific Islander/<br>Hawaiian Native | 6/9797 (0.06%) | 0/2190 (0%) | 6/6255 (0.1%) | 0/1352 (0%) | 0.20 |
| Declined/Other | 1829/9797<br>(18.7%) | 476/2190<br>(21.7%) | 1117/6255<br>(17.9%) | 236/1352<br>(17.5%) | 0.69 |
| **Test Results** |  |  |  |  |  |
| Cancer &<br>Cardiovascular<br>Disease Variants<sup>g</sup> | 813/9797 (8.3%) | 177/2190 (8.1%) | 537/6255 (8.6%) | 99/1352 (7.3%) | ... |
| Tier 1 Pathogenic<br>Variants (HBOC,<br>Lynch Syndrome, &<br>FH)<sup>h</sup> | 182/9797 (1.9%) | 42/2190 (1.9%) | 114/6255 (1.8%) | 26/1352 (1.9%) | ... |
| HBOC | 116/9797 (1.2%) | 27/2190 (1.2%) | 73/6255 (1.2%) | 16/1352 (1.2%) | ... |
| Lynch Syndrome | 38/9797 (0.39%) | 3/2190 (0.14%) | 28/6255 (0.45%) | 7/1352 (0.52%) | ... |
| FH | 29/9797 (0.30%) | 12/2190 (0.55%) | 13/6255 (0.21%) | 4/1352 (0.30%) | ... |
| Non-Tier 1<br>Pathogenic Variants | 631/9797 (6.4%) | 135/2190 (6.2%) | 423/6255 (6.8%) | 73/1352 (5.4%) | ... |
| Pharmacogenomic<br>Variants<sup>i</sup> | 9607/9797<br>(98.1%) | 2154/2190<br>(98.4%) | 6128/6255<br>(98.0%) | 1325/1352<br>(98.0%) | ... |
| Actionable<br>Pharmacogenomic<br>Variants | 9599/9607<br>(99.9%) | 2150/2154<br>(99.8%) | 6124/6128<br>(99.9%) | 1325/1325<br>(100%) | ... |

**Table 1. DNA10K Primary Care Physician Mediated Genetic Screening Results (continued)**
| Variable | Overall<br>No./Total (%) or<br>Median (IQR) | FM<br>No./Total (%)<br>or Median<br>(IQR) | IM<br>No./Total (%)<br>or Median<br>(IQR) | OBGYN<br>No./Total (%)<br>or Median<br>(IQR) | p-value<sup>a</sup> |
| --- | --- | --- | --- | --- | --- |
| **Test Results per PCP** |  |  |  |  |  |
| **Patients who Completed Test per PCP** | 68.5 (25.5-126) | 62.5 (33-120) | 66 (22.5-138) | 75 (40-118.2) | ... |
| **Cancer & Cardiovascular Disease Variants per PCP<sup>g</sup>** | 7.5 (4.9-10.2) | 6.8 (3.7-9.1) | 7.8 (5.6-10.8) | 7.4 (5.5-8.8) | 0.40 |
| **Tier 1 Pathogenic Variants per PCP (HBOC, Lynch Syndrome, & FH)<sup>e</sup>** | 1.4 (0-2.5) | 1.5 (0-2.8) | 1.4 (0-2.5) | 1.3 (0-2.4) | 0.97 |
| **HBOC per PCP** | 0.8 (0-1.5) | 0.7 (0-1.5) | 0.7 (0-1.5) | 1.0 (0-1.8) | 0.95 |
| **Lynch Syndrome per PCP** | 0 (0-0.4) | 0 (0) | 0 (0) | 0 (0-1) | 0.12 |
| **FH per PCP** | 0 (0) | 0 (0) | 0 (0) | 0 (0-0.2) | 0.08 |
| **Non-Tier 1 Pathogenic Variants per PCP** | 5.8 (3.5-7.5) | 5.0 (2.1-7.0) | 6.1 (3.8-8.6) | 5.8 (3.7-6.8) | 0.28 |
| **Actionable Pharmacogenomic Variants per PCP<sup>i</sup>** | 98.3 (97.2-100) | 98.6 (97.5-100) | 98.2 (96.7-99.6) | 97.9 (97.3-100) | 0.39 |

Abbreviations: FH, Familial Hypercholesterolemia; FM, Family Medicine; HBOC, Hereditary Breast and Ovarian Cancer; IM, Internal Medicine; IQR, Interquartile Range; OBGYN, Obstetrics/Gynecology; PCP, Primary Care Physician; SD, Standard Deviation.
<sup>a</sup> P values are for the Kruskal-Wallis test unless otherwise indicated; $P < .05$
<sup>b</sup> ANOVA with post-hoc Tukey HSD Test.
<sup>c</sup> FM vs. IM, $p < 0.05$ .
<sup>d</sup> IM vs. OBGYN, $p < 0.05$ .
<sup>e</sup> OBGYN vs. FM, $p < 0.05$ .
<sup>f</sup> Wilcoxon Rank Sum Test comparing females across FM and IM only.
<sup>g</sup> The Color Genomics, Inc. next generation sequencing panel includes the following genes:
Cancer Risk (30 genes): *APC, ATM, BAP1, BARD1, BMPR1A, BRCA1, BRCA2, BRIP1, CDH1, CDK4, CDKN2A* (p14ARF, p16INK4a), *CHEK2, EPCAM, GREM1, MITF, MLH1, MSH2, MSH6, MUTYH, NBN, PALB2, PMS2, POLD1, POLE, PTEN, RAD51C, RAD51D, SMAD4, STK11, TP53*.

## Discussion

In this study, the engagement of an integrated health system in the implementation of a genetic screening program enabled PCPs to identify cancer and cardiovascular disease variants in more than 8% of patients and at least one CDC Tier 1 variant in nearly 2% of tested patients. Limitations included the lack of detailed family and medical histories to assess the proportion of patients with actionable variants who met evidence-based criteria for genetic testing. Additionally, there was not sufficient follow-up time to assess the effects of testing on preventive screenings such as mammography, colonoscopy, cardiometabolic testing, and genetics referrals. However, results suggest population-based genetic screenings mediated and personalized by PCPs have major potential to impact population health outcomes if identified actionable variants lead to preventive interventions with established clinical utility

## Data Availability

Dr. David had full access to all of the data in the study and takes responsibility for the integrity of the data and the accuracy of the data analysis.

## Author Contributions

*Concept and Design*: David, Dunnenberger, Ali, Zimmer, Lemke, Hulick

*Acquisition, analysis, or interpretation of data*: David, Ali, Matsil

*Drafting of the manuscript*: David, Ali

*Revision of the manuscript*: David, Dunnenberger, Ali, Matsil, Zimmer, Lemke, Hulick

*Statistical analysis*: Singh, Matsil, Ali

*Supervision:* David

## Role of the Funder/Supporter

The funders did not contribute to the design, conduct, collection of data, analysis or interpretation of the data, or influence or have editorial control on the content or review of the manuscript or decision to submit the manuscript for pre-print publication.

## Additional Contributions

We wish to acknowledge the leadership of John Revis, MD, Janardan Khandekar, MD, and Justin Brueck, MHA at NorthShore University HealthSystem (Evanston, IL), NorthShore Health Information Technology and EPIC Ambulatory Team for information technology EMR integration work, Alicia Zhou, PhD at Color Genomics Inc. (Burlingame, CA) for support, and EPIC Systems™ (Verona, WI) for assistance with delivery of genomic services and EMR integration.

